# COVID-19 infections in day care centres in Germany: Social and organisational determinants of infections in children and staff in the second and third wave of the pandemic

**DOI:** 10.1101/2021.06.07.21257958

**Authors:** Franz Neuberger, Mariana Grgic, Svenja Diefenbacher, Florian Spensberger, Ann-Sophie Lehfeld, Udo Buchholz, Walter Haas, Bernhard Kalicki, Susanne Kuger

## Abstract

**Background:** During the SARS-CoV-2 pandemic, German early childhood education and care (ECEC) centres organised childrens attendance variably (i.e., reduced opening hours, emergency support for few children only or full close-down). Further, protection and hygiene measures like fixed children/staff groups, ventilation and surface disinfection were introduced among ECEC centres. To inform or modify public health measures in ECEC, we investigate the occurrence of SARS-CoV-2 infections among children and staff of ECEC centres in light of social determinants (socioeconomic status of the children) and recommended structural and hygiene measures. We focus on the question if the relevant factors differ between the 2nd (when no variant of concern (VOC) circulated) and the 3rd wave (when VOC B.1.1.7 (Alpha) predominated).

**Methods:** Based on panel data from a weekly online survey of ECEC centre managers (calendar week 36/2020 to 22/2021, ongoing) including approx. 8500 centres, we estimate the number of SARS-CoV-2 infections in children and staff using random-effect-within-between (REWB) panel models for count data in the 2nd and 3rd wave.

**Results:** Centres with a high proportion of children with low socioeconomic status (SES) have a higher risk of infections in staff and children. Strict contact restrictions between groups like fixed group assignments among children and fixed staff assignments to groups prevent infections. Both effects tend to be stronger in the 3rd wave.

**Contribution:** ECEC centres with a large proportion of children from a low SES background and lack of using fixed child/staff cohorts experience higher COVID-19 rates. Centres should be supported in maintaining recommended measures over the long run. Preventive measures such as vaccination of staff should be prioritised in centres with large proportions of low SES children.

## Background

Germany faced three pandemic COVID-19 waves so far. While the 1st wave in spring 2020 was followed by a phase of low incidence during summer 2020, the 2nd wave started approximately in CW 40/2020 and lasted until the first weeks in 2021. The 3rd wave followed on foot and was characterised by a parallel rise of the proportion of specimens diagnosed as the VOC B.1.1.7 (“British variant” resp. “Alpha”). To curb incidences, the German government ordered national lockdowns and reduced the number of children in early childhood education and care (ECEC) centres to reduce the number of contacts [1–5].

### Childrens Attendance in German ECEC Centres during the Corona Pandemic

With beginning of the 1st wave, a “strict” lockdown was introduced during which only children of parents providing essential services (e.g. physicians or food vendors) and children in need of child welfare services (e.g. cases of maltreatment) could attend ECEC centres [5]. During the 2nd wave, a 2nd lockdown was installed from CW 51/2020 until CW 04/2021. Throughout this lockdown, all children could attend ECEC in principle. However, most federal states appealed to parents to keep their children at home if possible [1–4]. During the 3rd pandemic wave (since CW 05/2021), implementation of ECEC closure was largely dependent on the incidence of individual counties. Hence, ECEC attendance regulations differed across regional meso and micro levels.

### Transmission of SARS-CoV-2 involving Young Children: unclear role of Preventive and Hygiene Measures

Before circulation of VOC B.1.1.7 (i.e., prior to the 3rd wave), children 1-11 years old were under-represented among COVID-19 cases compared to their proportion in the general population and particularly under-represented among cases experiencing severe outcomes, such as hospitalisation, requiring respiratory support or death [6]. Two systematic literature reviews conducted in 2020, i.e. before circulation of VOC B.1.1.7, concluded that they were less susceptible than adults [7, 8]. Data on infectiousness have shown equivocal results. In household studies children were rarely identified as primary cases [6] and gave rise to a lower (secondary) attack rate ((S)AR) of 7.9% (95% confidence interval (CI), 1.7%-16.8%) compared to that of adults (15%, 95% CI, 6.2%-27%) [9]. In German ECEC centres, children with COVID-19 infections have led to an average SAR of 1.7% and small outbreak sizes (average size: 3-4 cases) [10].

Transmission of SARS-CoV-2 occurs mainly through the respiratory route. Within the respiratory route, both short and long range transmissions are believed to play a role [11–13]. Exhaled aerosols can float in the air for hours [14], and half life of viable virus in small particles is estimated as approximately one hour [15]. Once certain boundary data, such as room size, duration of exposure, number of persons exposed and type and degree of ventilation are known, it has been possible to predict the attack rate of outbreaks [16]. The role of other transmission routes, e.g. contact transmission, remains controversial. Ferretti estimates that 10% of transmissions may be due to “environmental” factors, i.e. contact transmission [17]. Meyerowitz concludes that there is currently no conclusive evidence for fomite or direct contact transmission of SARS-CoV-2 in humans [13].

Conversely, at least among households, ventilation was shown to be protective for secondary infections [18]. These findings have led to the recommendations to keep a minimum distance of 1.5 m to other persons, wear masks and ventilate rooms where several persons are present at the same time. In principle, these recommended behaviours provide protection in the context of ECEC centres as well. However, as most transmission studies have been conducted among adults, the evidence base for preventive recommendations among children is largely unexplored. For children at preschool age, it cannot be expected to keep a distance of 1.5 m to peers or staff, nor to wear masks. Recommended measures for ECEC centres have thus focused on organisational changes, e.g. the change of group concepts as well as infection control / hygiene recommendations for staff and parents.

Before the pandemic, the following (pedagogical) group concepts typically existed in German ECEC settings: (i) fixed groups, (ii) open concept (children can freely choose and switch between rooms and peers), and (iii) partly open concept, e.g. a fixed group in the morning and free roaming in the afternoon. These concepts leave all options open how staff is assigned to groups. An important organisational change in ECEC centres was the recommendation to switch not only to fixed groups, but to also keep pedagogical staff of a given group constant (fixed staff assignment to a particular group). In addition, infection control and hygiene recommendations included regular ventilation of rooms and regular disinfection of surfaces [19].

### Project Description

The German Youth Institute (“Deutsches Jugendinstitut”; DJI) and the Robert Koch Institute (the national public health institute; RKI) joined forces to monitor the situation of pre-school children during the pandemic in the so called “Corona-KiTa-Study”. In one of the project modules, the DJI established a novel surveillance system drawing information directly from ECEC centres (the so-called “KiTa-Register” which translates into “ECEC centre registry”). It has been set up to monitor the re-opening and closing of ECEC centres, the rate of children attending settings and the staff in attendance during the pandemic. Furthermore, information on COVID-19 infections in ECEC centres and the implementation of infection control and hygiene measures has been collected.

### Research Questions

We put a focus on the questions (1) to understand relevant determinants, risk and protective factors for COVID-19 occurrence in ECEC centres, (2) whether factors associated with infections differ between children and staff, and (3) as the 3rd wave is largely driven by the mutated VOC B.1.1.7 [20] with possibly different epidemiological properties if the relevant factors differ between the 2nd and 3rd wave.

## Methods

### Data

To investigate possibly predictive and protective factors of COVID-19 infections in ECEC centres, this paper uses the panel data set of ECEC centres in Germany within the Corona-KiTa-Study (ECEC centre registry). From CW 36 in August 2020 managers of all ECEC centres in Germany were asked to fill out a weekly questionnaire. Until CW 22/2021, 8,500 of roughly 54,000 ECEC centre managers participated at least once with an average number of 29.5 reported weeks. The survey comprises a baseline questionnaire distributed at registration to collect basic time-constant information such as the provider type, the centres proportion of children from households with low socioeconomic status (SES) and the group concept prior to the pandemic. The subsequent weekly questionnaire collects information about the current week and contains time-varying variables such as the number and age of children currently attending the ECEC centre, the number of staff working at the ECEC centre in general as well as in the current week, the currently applied group concept, if staff was assigned firmly to groups (fixed staff assignment (only asked if ECEC currently use a fixed or partly open group concept)), application of certain hygiene measures, as well as the number of children, staff and parents who are tested positive for COVID-19.

### Dependent variables

We use the number of reported infections per week as dependent variable. To measure infections in staff and children, ECEC centre managers were asked if they had any new laboratory confirmed cases of COVID-19 in children or staff. Infections were reported for staff members and for children separately. For data protection reasons, detailed information on infections in staff was only asked in ECEC centres with at least 7 staff members (which applies to 97% of our sample). The serial interval of COVID-19 (i.e., the average interval from the onset of illness in an infectious / case to the onset of illness of a case infected by that case) is estimated to have a duration of 5 days [21, 22]. After laboratory diagnosis, 1-3 days may pass until the result of a test is available at the county health department [23]. Hence, we included the variable “number of infections” with a lead of one week in our models and estimated the rate of infection in CW X+1 with data from CW X. We do not analyse infections in parents, as the link to the ECEC centre is not necessarily given here.

### Independent variables

As possible predictors for the number of reported infections, we use variables which are either time-constant or time-variant.

#### Time-constant variables

##### Type of provider

The type of provider (public, private for-profit, ecclesiastical or other non-profit) of the ECEC centres is included, since it might be associated with the (systematic) implementation of particular hygiene measures.

##### Socioeconomic status

COVID-19 infections are known to follow a social gradient [24, 25]. To control for social composition, ECEC managers were asked to estimate the proportion of children with low socioeconomic status (SES) on a 4 point Likert scale (i.e., below 10% children with low SES background, 11% to 30%, 31% to 60%, or above 60%, respectively).

##### Group concept prior to the pandemic

To grasp differences in the set-up of the institutions which might make it difficult to implement certain measures such as e.g. group separation, we include the type of pedagogical group concept before the pandemic in our model.

We include these variables as time-constant dummy variables in the models.

#### Time-varying variables

##### Currently applied group concept

Managers indicated which group concept was currently in use (i.e., fixed, open, partly open, see above). We analysed the currently applied group concept as this may facilitate close contact to more or less children or staff in the ECEC centres. We distinguished between the three categories “open group concept”, i.e. attendants could mix freely, “partly open” and “fixed”, i.e. strict assignment of children to only one group.

##### Infection control and hygiene measures

Managers specified which other measures they took: (1) Regular ventilation of the rooms, (2) regular surface disinfection (e.g., furniture surfaces, door handles or toys), and (3) a fixed staff assignment to groups (only asked for fixed or partly open group concepts).

#### Control variables

##### 7-day incidence on county level

Laboratory confirmed COVID-19 cases are notified to the local health authority (LHA) in accordance with the German Protection against Infection Act (IfSG)^1^. The LHA transmits reported cases via the respective federal state health authority to the RKI. The 7-day incidence includes the number of newly reported cases within seven days per 100,000 population.

##### Number of Children

Managers indicated per week how many children aged 0-2, 3-5, and 6 years and older attended the ECEC centres. These numbers may change due to (perhaps only regionally observed) holidays as well as regional outbreaks and measures taken by the federal state or county.

We use the latter two variables, 7-day incidence on county level and number of children attending, to control for possible exposure to the virus.

### Statistical analyses

Our data provide various information about ECEC centres, namely on *time-constant* variables related to the centre from the baseline questionnaire, on average differences in the time-varying variables *between* ECEC centres and on changes *within* a centre as well. Since the Poisson distribution is known to approximate incidence counts from a wide variety of underlying processes [26], we use a random-effect panel poisson model for count data with demeaned data to approximate incidence counts (see formula 1). That allows us to estimate the effects of time-constant variables, between-unit differences and within-unit changes on the incidence count at the same time [27, 28].

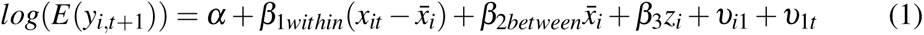

We specify our model with a leading y variable (t+1). *β*_1_ estimates within-units effects, hence what happens in the next weeks after a unit e.g. decides to change its group concept to open. *β*_2_ estimates between-units effects, hence the effect of e.g. having an open group concept all the weeks under study. *β*_3_ estimates effects of time-constant variables, e.g. of applying an open group concept before the pandemic. *υ*_*i*1_ and *υ*_1*t*_ are unit- and time-fixed effects. Exponential coefficients could be interpreted as incidence rate ratios, hence how much the expected count changes multiplicatively when x increases by one.

As the occurrence of COVID-19 infections varies by time and region, we include a county‘s 7-day incidence as (log) exposure A [29] with a regression coefficient constrained to 1, allowing the model to represent rates instead of counts. This is equivalent to standardizing the dependent variable with the offset variable (see formula 2, equivalent with formula 3).

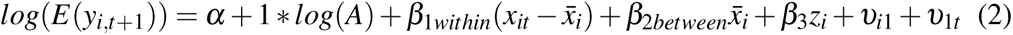

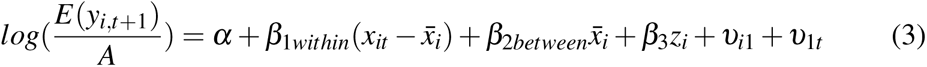

By doing so, we include the assumption that an ECEC centre in a county with twice as many infections also reports twice as many cases in the ECEC centre. Since the likelihood of an occurrence of a COVID-19 infection does not only depend on regional conditions, but is also strongly dependent on the number of persons in the respective facilities, we add within- and between-effects for the number of children of all age groups (0-2, 3-6, 7 plus) in our model. As we tend to refrain from interpreting these variables directly, they are included in the model as mere controls and are only shown in the appendix tables.

## 1 Results

### COVID-19 infections in ECEC centres over time

Figure 1a shows the development of COVID-19 infections in children and staff between CW 36/2020 and CW 22/2021. The gray area in Figure 1 demarcates the 2nd wave (including CW 04 in 2021), the remaining white area (CW 05-22 in 2021) demarcates the 3rd wave. Our wave definitions do not exclude weeks with low incidence rates as we want to measure within and between effects, hence we need to include the lead time before incidences rise.

**Figure 1:**
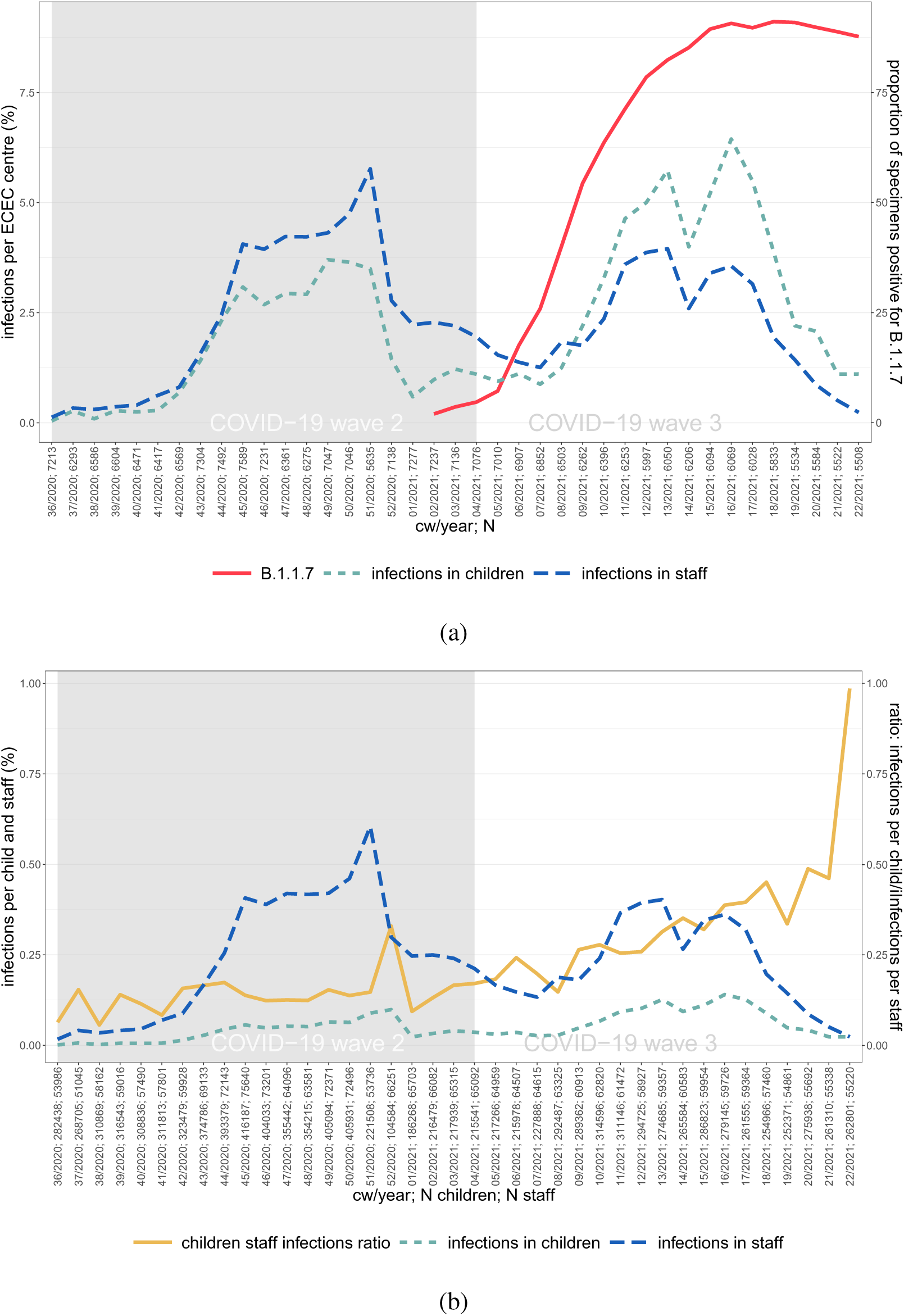
Infections in children and staff. (a) Average number of infections (n) in children and staff in ECEC centres per week, (n/N per week) and proportion of specimens in German laboratories positive for B.1.1.7 (%) (b) Average number of Infections in children and in staff per week *Source: survey data (ECEC centre registry) collected by the German Youth Institute, own calculations. RKI Data on B.1.1.7; Robert Koch-Institut (2021): Bericht zu Virusvarianten von SARS-CoV-2 in Deutschland (April)*.

In the 2nd wave of the pandemic, the weekly number of infections in staff per ECEC centre exceeds the number of infections in children per ECEC centre. From CW 9/2021, the two lines cross and the number of infected children per ECEC centre surpasses that of staff. To account for the number of children and staff attending week by week, Figure 1b shows the proportion of infections in ECEC staff and children, respectively, as well as the ratio of the proportion of infections among children relative to the one in staff, the latter could be interpreted as a rate ratio. Overall, the rate of infections in staff is higher compared to that of children, but the ratio clearly increases in the 3rd wave, and further rises when infections go back in recent weeks.

### Attendance and protection and hygiene measures in ECEC centres over time

In the following, we briefly discuss the development of our time-varying indicators. As we specify our model with a 1 week lead, explanatory variables are only shown up to CW 21/2021. Respective figures and tables are included in the appendix. Regarding *childrens attendance*, prior to the 2nd wave, the average ECEC centre each week cared for roughly 12 children aged 0 to 2 years, for 45 children aged 3 to 6 years and for 3-4 children aged 7 years or older (see Appendix Figures A3a, A3b and Tables 3, 4 for details). The latter number might seem low, but our data collection excludes all ECEC centres with after-school childcare that do not provide ECEC services for younger children. Overall, we found only little variation in the number of children attending child care in the weeks 36 to 50 in 2020. We observe a steep drop in childrens attendance in CW 50 to 52 in 2020, as the German federal government ordered a nationwide lock-down in which most federal states appealed to parents to keep their children at home if possible [1–4]. Roughly half of the children returned after Christmas, while the 2nd half returned in CW 8/2021, followed by two minor increases in CW 15 and 20/2021.

The share of ECEC centres applying a fixed group concept (see Appendix Figures A4a, A4b and Tables A3, A4) increased from 62% in CW 36/2020 to 80% in CW 21/2021. At the same time ECEC centres with a partly or fully open concept decreased from 29% to 15% and 8% to 5%, respectively.

Considering *infection control and hygiene measures*, regular ventilation was conducted in most ECEC settings during the whole period and decreased only slightly from 100% of institutions in CW 36/2020 to 97% in CW 21/2021 (see Appendix Figures A5a, A5b and Table A3, A4 in the Appendix). The vast majority of ECEC centres indicated to have disinfected surfaces regularly during the whole time under study, with only a small drop from 92% in CW 36/2020 to 88% in CW 21/2021. Only 63% of ECEC centre managers reported that their staff was assigned firmly to a fixed or partly open group in CW 36/2020, but this share increased to 69% in CW 21/2021. Despite these relatively small changes over time, we found considerable within-variance, above all in the latter variable. We observed simultaneous changes in different directions, as some started and others stopped infection control and hygiene measures in the same week, leading to a low change in overall percentages.

### Factors associated with COVID-19 infections in ECEC centres

Figure 2 shows results from the REWB models. It contains coeffients from models analysing the number of infections within different time frames (i.e. 2nd and 3rd wave of the pandemic) in both staff and children, hence four coefficients per explaining variable. The coeffients are shows as incidence rate ratios. The point estimates are marked with a dot (for infections in staff, Model 1 and 3) or a square (for infections in children, Model 2 and 4). Horizontal lines indicate the confidence bounds (95%), where the linetype marks the wave, i.e. 2nd wave (solid, Model 1 and 2) and 3rd wave (dashed line, Models 3 and 4). Models 1 and 2 cover a 21-week time frame from CW 36/2020 to CW 04/2021 (i.e., 2nd wave, the grey area in Figure 1), while Models 3 and 4 cover a 17-week time frame from CW 05/2021 to CW 22/2021 (i.e., 3rd wave, the white area in Figure 1). Significant coefficients (*z* < .1) are printed in opague, non significant coeffients are printed in transparent colours. Signficant levels are further printed as text in the corresponding colours.

**Figure 2:**
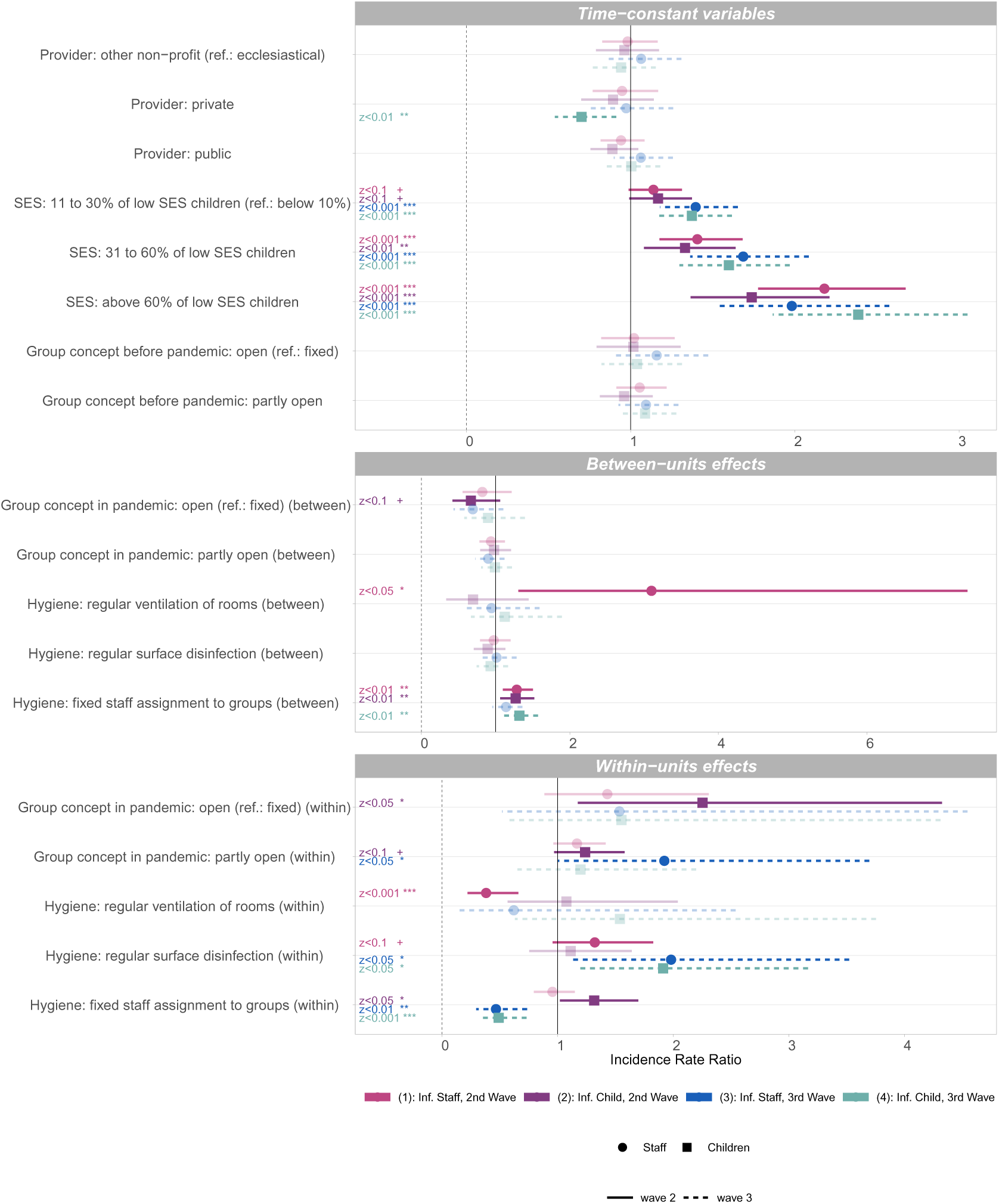
Incidence rate ratios of predictors for infections in staff and children in 2nd (Model 1,2) and 3rd wave (Model 3,4) of the pandemic. \*\*\**z <* 0.001; \*\**z* < 0.01; \**z* < 0.05; +*z* < 0.1. Source: Survey data collected by the German Youth Institute (ECEC centre registry). Second wave assumed to last from calendar week (CW) 36/2020-04/2021, and third wave from CW 05/2021-22/2021. REWB poisson model with two-way fixed effects, offset for county incidence (data collected by the Robert Koch-Institute), dependent variable with 1 week lead. Coefficients are displayed as incidence rate ratio, confidence bounds (95%) as bars, effects that do not reach the threshold of *z <* 0.1 are transparent, significant effects are shown as opaque. Controlled for number of children in different age groups (within and between effects), see Appendix Table A1 for full model, own calculations.

#### Results for time-constant variables

We do not find significant differences for different *types of providers*, except a negative effect of private providers on infections in children in wave 3 (Model 4). The *proportion of low SES children* is found to be a significant predictor for the rate of infections in staff as well as in children in all four models for all categories of SES. The higher the proportions of children from a low SES background, the higher the infection rate. For 11% to 30% of low SES children (compared to less than 10% low SES children), effects are only significant at the 10% significance level (^†^*p <* 0.1) in Models 1 and 2 (i.e., 2nd wave). Effect sizes of SES tend to be larger in wave 3 compared to wave 2 for both infections in staff and children, with one exception for staff regarding the effect of the largest proportion of children with low SES background, which is larger in wave 2 than in wave 3. The variable *group concept prior to the pandemic* did not yield any significant effects.

#### Results for time-varying variables: Between-units effects

Between-units effects compare averages across CWs between ECEC centres and are prone to endogeneity, that is, a positive effect (indicating more infections) of implementing a protection measure could stem from the fact that centres in counties with very low COVID-19 incidences tend to implement protection measures to a lower extent than centres in counties with higher incidences.

We do not find significant between-units effects regarding the *currently applied group concept*, with the exception of a negative effect of open group concept at the 10% level for infections in children in the 2nd wave (Model 2), indicating that, controlling for the factors in the model, ECEC centres with low infection rates tend to apply open group concepts more often.

We find a significant positive between-units effect for *regular ventilation* for infections in staff in wave 2 (Model 1). We find no between-unit effects of regular surface disinfection. We find positive between-units effects for *fixed group assignment of staff* (Models 1, 2, and 4, i.e. wave 2 for staff and children and wave 3 for children). The positive effects indicate that, controlling for the factors in the model, ECEC centres that report to adhere to the recommendation to ventilate rooms more frequently or have a fixed staff assignment to groups more often on average (i.e., implement this measures in more weeks) also report higher average infection rates in the respective settings.

#### Results for time-varying variables: Within-units effects

Next, we describe time-varying coefficients for within-units effects which estimate the effect of a within-unit changepage For example, in the case of the variable group concepts, we estimate what happens if ECEC centres switch from a fixed to an open group concept between weeks. Results thus imply a temporal association of events within an ECEC centre.

Considering the *group concept*, we find significant within-units effects. Switching to an open concept (from a fixed concept) is associated with a significant increase in the number of infections among children in the 2nd wave (Model 2). Switching to a partly open concept (from a fixed concept) is associated with a significant increase of infections in children in the 2nd wave (Model 2), and more so, with increasing infections in staff in the 3rd wave (Model 3).

Starting the implementation of *regular room ventilation* is significantly associated with fewer infections in staff in wave two (Model 1), but has no significant effects on the infection rates in any other Model (Models 2, 3 and 4).

Implementing *regular disinfection of surfaces* from one week to the next is associated with a significant increase of infections among staff and children in the 3rd wave (Model 3 and 4), and less so, with increasing infections among staff in the 2nd wave (Model 1, 10 % level). Further analysis showed that this effect is likely to stem from ECEC centres which started disinfection when local county incidences were already on the rise. Centres that reported the implementation of surface disinfection have a lower county average COVID-19 incidence before they start, and a higher county average after having started with disinfection. Hence, we assume that the start of surface disinfection was a reaction to locally rising incidence rates.

Implementing *fixed group assignment of staff* is associated with significantly fewer infections in both staff and children in wave 3 (Model 3 and 4). In the 2nd wave models, we found no effect for infections in staff, but a significant positive effect for infections in children (Model 2). The small positive effect in Model 2 might be again read as anticipation, hence ECEC centres start with fixed staff assignments to groups in the face of rising incidence rates, presumably reacting to regional orders for implementation which are linked to rising incidence rates.

To challenge our results, we additionally ran a variety of models as robustness checks, i.e., several models with protective measures only (Appendix Tables A5-A8, models that include the offset as variable (Appendix Table A9) and models with alternative wave cut-off points (Appendix Tables A10 and A11). All tests by and large confirm the above results in relation to the sign and the effect strength and can be found in appendix tables A5-A11. Considering protective measures, the very low number of within changes, especially in ventilation and disinfection, leads to a strong dependence on few observations only and the effects should therefore only be interpreted with caution.

We further tested if our data fits the poisson specification by analyzing the conditional mean and variance of the outcome variables in our models, finding underdispersion in all our models. Underdispersion leads to overestimated standard errors in poisson models [30]. As we prefer to not correct for that bias, e.g. by using a quasipoisson model, (which would lead to smaller confidence bounds), our significance levels could be characterised as conservative. This approach seems appropriate, especially in view of the fact that our data does not represent a true random selection. We tried alternative specifications for underdispersed data, i.e. using a generalised poisson distribution (see Appendix Table A12) that by and large confirms our results. Notable exceptions are the within-effect of fixed staff assignment for infections in children in wave 3 (Model 4) which is smaller and insignificant as well as the within effects of surface disinfection, which are insignficant when a generalised poisson distribution is assumed.

## Discussion

Our study investigated factors associated with infections in staff and children in ECEC centres in Germany during the 2nd and 3rd wave of pandemic. We found ECEC centres with a larger proportion of children from a low SES background to have the highest risk of infections, and we found this effect to be increasing in wave 3 compared to wave 2.

The change of *group concepts* towards a more restrictive group separation was one of the essential recommended or ordered measures since the beginning of the pandemic. A large proportion of ECEC centres in our sample followed this recommendation, especially in the 3rd wave (Appendix Figure A4). The significant associations of switching from a fixed to a open or partly open concept shows that failure to follow these recommendations puts children and staff health at risk.

It is well recognised that the main route of SARS-CoV-2 transmission is through the respiratory mode, and that airborne transmission is of major importance. Our findings tend to confirm this for ECEC settings. In our data, regular ventilation had a significant protective effect on infections in one of the four models, specifically on staff in the 2nd wave (model 1). Overall, implementation of *regular ventilation* as a protective measure in ECEC centres was quite complete with only very little variance. Hence, insignificance is probably due to very low case numbers resulting in low overall variance. Nevertheless, even if regular ventilation was widely implemented during the whole period under study, we observed a small, but constant decline in implementation.

In line with controversial findings regarding the role of contact or fomite transmission, we find significant positive within result for regular surface disinfection only in our models. We have no evidence that surface disinfection prevents COVID-19 cases, but it is unlikely that it creates cases. We believe that the most likely explanation is that ECEC managers anticipate or react to local outbreaks or increasing COVID-19 incidences with implementing hygiene measures, such as disinfection of surfaces. Hence, we do not know to which extent our results are biased due to anticipation.

In the 3rd wave, some ECEC centres switched to a more flexible staff assignment to groups and changed their group concept between CW 07 and 08 (see Appendix Figure A5b, A4b), presumably due to a shortage of staff. This was associated with an increase of infections in the 3rd wave. Thus, our results confirm that it should be beneficial for ECEC centres to maintain fixed staff assignment in order to prevent infections, particularly among staff.

Although it is unknown which infections in the data are due to a VOC, the 3rd wave was dominated increasingly by the VOC B.1.1.7, which is associated with increased transmissibility [31]. This again is in line with our finding that the effects of SES are stronger in wave 3, and also with our finding of stronger effects of fixed staff assignment in the 3rd wave. In this situation of VOC predominance, the implementation of contact restrictions between groups of children seem to gain priority to keep ECEC services open over the long run.

We acknowledge as a general limitation that a managers decision to implement specific measures in their ECEC centre (and the according report in our questionnaires) is not always followed and translated into every-day practice by all staff members. Further, some measures are conceptually similar, e.g. a fixed group concept (with consequent separation of children) and a fixed staff assignment which includes that also staff does not move between groups. As both show significant effects, this suggests that strict contact restrictions are likely to be one of the most effective protective measures, especially in the 3rd wave. It must be further mentioned that there is a certain chance that especially the results of surface disinfection and ventilation are driven by very few units within-changes (Appendix Figure A5b), as is further shown in the robustness section in the appendix (e.g., the within effect of surface disinfection is dependent on the wave definition, see appendix tables A10 and A11). This limitation does not hold true for the effect of fixed staff assignment on infections in staff, which remains significant in wave 3 in all robustness checks. We therefore strongly support the recommendation to keep up fixed staff assignment in all ECEC centres wherever possible.

## Conclusions

The result that about one third of the centres have not implemented the recommended fixed staff assignment is likely associated with well-known structural problems of many ECEC centres in Germany, which have to deal with limited staffing overall and within group settings and a shortage of professional staff recruitable on the labour market for years [32]. Because of this, the measure was only recommended but not prescribed by law. In order to better prepare ECEC centres for such exceptional situations in the future, it is essential to finally eliminate the staff shortage that was already prevalent before the pandemic.

In summary, our study suggest that the COVID-19 pandemic affects ECEC centres particularly when they were attended from children with low SES. The social gradient of COVID-19 does not stop at the ECEC centre’s door, indicating that children, families and staff in corresponding centres need special support. Although many ECEC centres are grappling with staffing difficulties it is important to maintain – to the best possible degree – fixed group assignments among children and fixed staff assignments to groups. In addition, generous and frequent ventilation may aid in preventing infections. As vaccinations become increasingly available and booster vaccinations will probably become necessary in the light of further SARS-CoV-2 variants, particularly staff of ECEC centres with a large proportion of children from a low SES background should be prioritised.

## Data Availability

Anonymized data will be passed on to external researchers after the project has been finalized (Dec. 2021), provided that they are used for the purpose of scientific secondary and subsequent use and under the conditions of the DJI Research Data Centre (incl. deletion periods, thematic limitation, citation, etc.). The anonymised survey data and research results are stored for 10 years as proof of good scientific practice and in accordance with the funding regulations.

## List of abbreviations

CW: Calendar week
DJI: German Youth Institute (Deutsches Jugendinstitut)
ECEC: Early childhood education and care
IfSG: German Protection against Infection Act (Infektionsschutzgesetz)
LHA: Local health authority
REWB: Random-effect-within-between
RKI: Robert Koch-Institute
SES: Socioeconomic status
VOC: Variant of concern

## APPENDIX COVID-19 infections in day care centres in Germany: Social and organisational determinants of infections in children and staff in the second and third wave of the pandemic

**Table A1:** Full Models from Figure 2 including effects for number of children. Incidence rate ratios (log) of predictors for infections in staff and children in 2nd (Model 1,2) and 3rd wave (Model 3,4)

|  | (1): Inf. Staff, 2nd Wave | (2): Inf. Child, 2nd Wave | (3): Inf. Staff, 3rd Wave | (4): Inf. Child, 3rd Wave |
| --- | --- | --- | --- | --- |
| Intercept | -10.77***<br>(0.45) | -9.55***<br>(0.40) | -9.78***<br>(0.30) | -9.69***<br>(0.29) |
| Provider: other non-profit (ref.: ecclesiastical) | -0.02<br>(0.09) | -0.04<br>(0.10) | 0.06<br>(0.11) | -0.06<br>(0.10) |
| Provider: private | -0.05<br>(0.11) | -0.11<br>(0.13) | -0.03<br>(0.13) | -0.35**<br>(0.13) |
| Provider: public | -0.06<br>(0.07) | -0.12<br>(0.08) | 0.06<br>(0.09) | 0.00<br>(0.08) |
| SES: 11 to 30% of low SES children (ref.: below 10%) | 0.13<sup>†</sup><br>(0.07) | 0.15<sup>†</sup><br>(0.08) | 0.33***<br>(0.09) | 0.32***<br>(0.08) |
| SES: 31 to 60% of low SES children | 0.34***<br>(0.09) | 0.29**<br>(0.11) | 0.52***<br>(0.11) | 0.47***<br>(0.11) |
| SES: above 60% of low SES children | 0.78***<br>(0.10) | 0.55***<br>(0.12) | 0.68***<br>(0.13) | 0.87***<br>(0.13) |
| Group concept before pandemic: open (ref.: fixed) | 0.02<br>(0.11) | 0.02<br>(0.13) | 0.15<br>(0.12) | 0.04<br>(0.12) |
| Group concept before pandemic: partly open | 0.05<br>(0.07) | -0.04<br>(0.08) | 0.09<br>(0.08) | 0.08<br>(0.08) |
| Group concept in pandemic: open (ref.: fixed) (between) | -0.20<br>(0.20) | -0.41<sup>†</sup><br>(0.24) | -0.37<br>(0.24) | -0.11<br>(0.22) |
| Group concept in pandemic: partly open (between) | -0.06<br>(0.09) | -0.02<br>(0.11) | -0.11<br>(0.11) | -0.01<br>(0.11) |
| Hygiene: regular ventilation of rooms (between) | 1.13*<br>(0.44) | -0.36<br>(0.37) | -0.06<br>(0.27) | 0.12<br>(0.27) |
| Hygiene: regular surface disinfection (between) | -0.03<br>(0.11) | -0.11<br>(0.12) | 0.01<br>(0.12) | -0.07<br>(0.11) |
| Hygiene: fixed staff assignment to groups (between) | 0.25**<br>(0.08) | 0.24**<br>(0.09) | 0.13<br>(0.09) | 0.28**<br>(0.09) |
| Group concept in pandemic: open (ref.: fixed) (within) | 0.36<br>(0.24) | 0.81*<br>(0.33) | 0.43<br>(0.55) | 0.44<br>(0.52) |
| Group concept in pandemic: partly open (within) | 0.15<br>(0.10) | 0.21<sup>†</sup><br>(0.12) | 0.65*<br>(0.33) | 0.18<br>(0.31) |
| Hygiene: regular ventilation of rooms (within) | -0.96***<br>(0.28) | 0.07<br>(0.33) | -0.48<br>(0.72) | 0.43<br>(0.46) |
| Hygiene: regular surface disinfection (within) | 0.28<sup>†</sup><br>(0.17) | 0.11<br>(0.20) | 0.68*<br>(0.29) | 0.65*<br>(0.26) |
| Hygiene: fixed staff assignment to groups (within) | -0.05<br>(0.09) | 0.27*<br>(0.13) | -0.76**<br>(0.23) | -0.71***<br>(0.20) |
| Nr. of children aged 0-2 (between) | 0.03***<br>(0.00) | 0.01***<br>(0.00) | 0.02***<br>(0.00) | 0.01*<br>(0.00) |
| Nr. of children aged 3-6 (between) | 0.01***<br>(0.00) | 0.01***<br>(0.00) | 0.01**<br>(0.00) | 0.01***<br>(0.00) |
| Nr. of children aged 7 and older (between) | 0.00<br>(0.00) | 0.01**<br>(0.00) | 0.00<br>(0.00) | 0.01<br>(0.00) |
| Nr. of children aged 0-2 (within) | -0.00<br>(0.00) | -0.02**<br>(0.01) | 0.03***<br>(0.01) | 0.00<br>(0.01) |
| Nr. of children aged 3-6 (within) | 0.01***<br>(0.00) | 0.01***<br>(0.00) | 0.01*<br>(0.00) | 0.01***<br>(0.00) |
| Nr. of children aged 7 and older (within) | 0.01<sup>†</sup><br>(0.00) | 0.01**<br>(0.00) | 0.00<br>(0.01) | 0.02***<br>(0.00) |
| AIC | 26728.74 | 19104.30 | 18125.15 | 23782.94 |
| Log Likelihood | -13337.37 | -9525.15 | -9035.58 | -11864.47 |
| Num. obs. | 114057 | 114057 | 87855 | 87855 |
| Num. groups: week | 21 | 21 | 16 | 16 |
| Num. groups: token | 7483 | 7483 | 6670 | 6670 |
| Var: week (Intercept) | 0.10 | 0.32 | 0.21 | 0.12 |
| Var: token (Intercept) | 2.14 | 2.42 | 2.58 | 2.93 |

\*\*\* $p < 0.001$ ; \*\* $p < 0.01$ ; \* $p < 0.05$ ; <sup>†</sup> $p < 0.1$ .. Source: Survey data collected by the German Youth Institute (ECEC centre registry). Second wave assumed to last from calendar week (CW) 36/2020-04/2021, and third wave from CW 05/2021-22/2021. REWB poisson model with two-way fixed effects, offset for county incidence (data collected by the Robert Koch-Institute), dependent variable with 1 week lead. Coefficients are displayed as log incidence rate ratio, standard errors in parentheses, own calculations.

**Figure A3:**
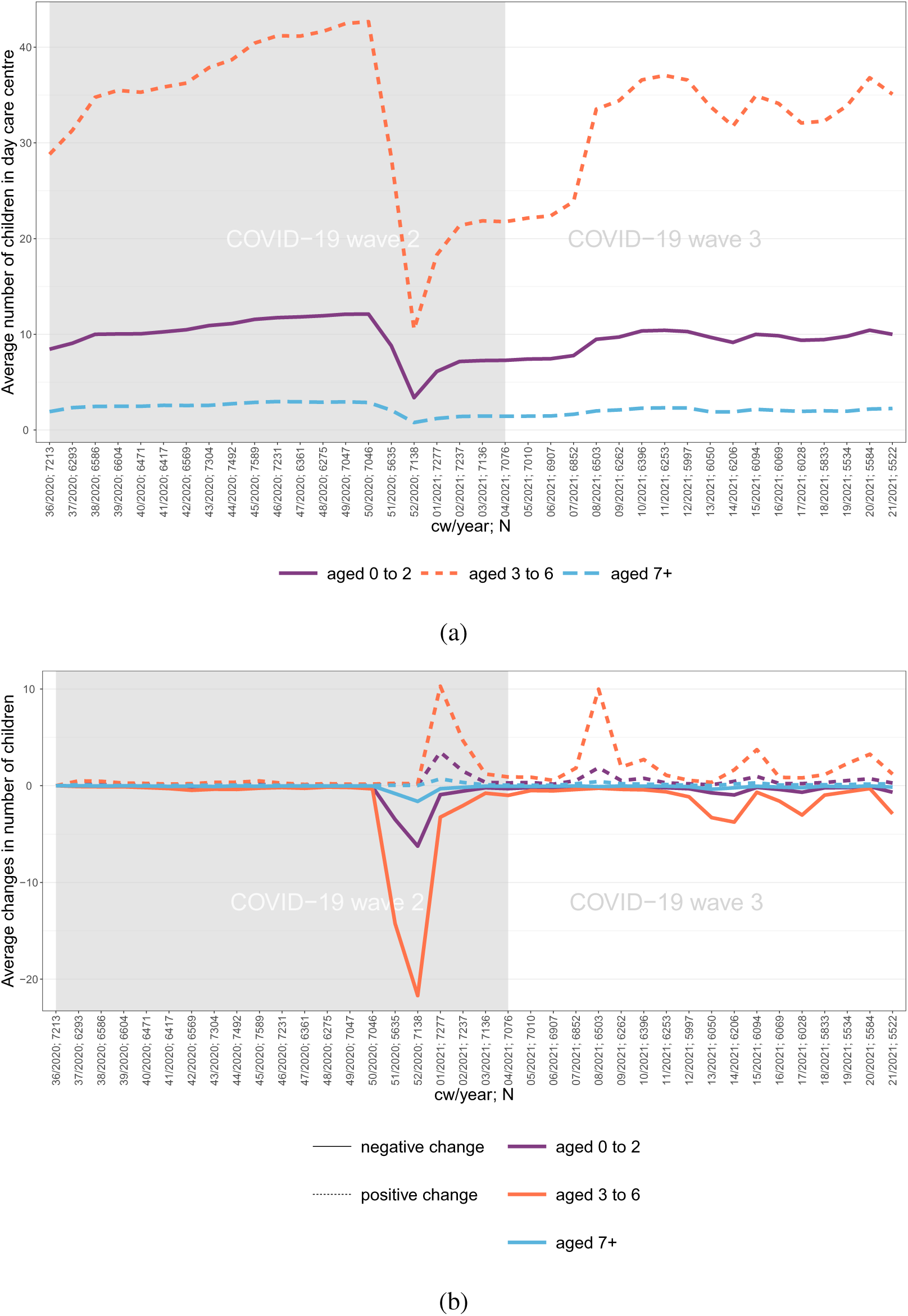
Number of children. (a) Average number of children per age group (n) visiting ECEC centres (N) per week, (n/N per week) (b) Average positive or negative change in number of children per age group (n) amongst ECEC centres (N) with positive or negative change, (n/N per week) *Source: Survey data collected by the German Youth Institute (ECEC centre registry), own calculations*.

**Figure A4:**
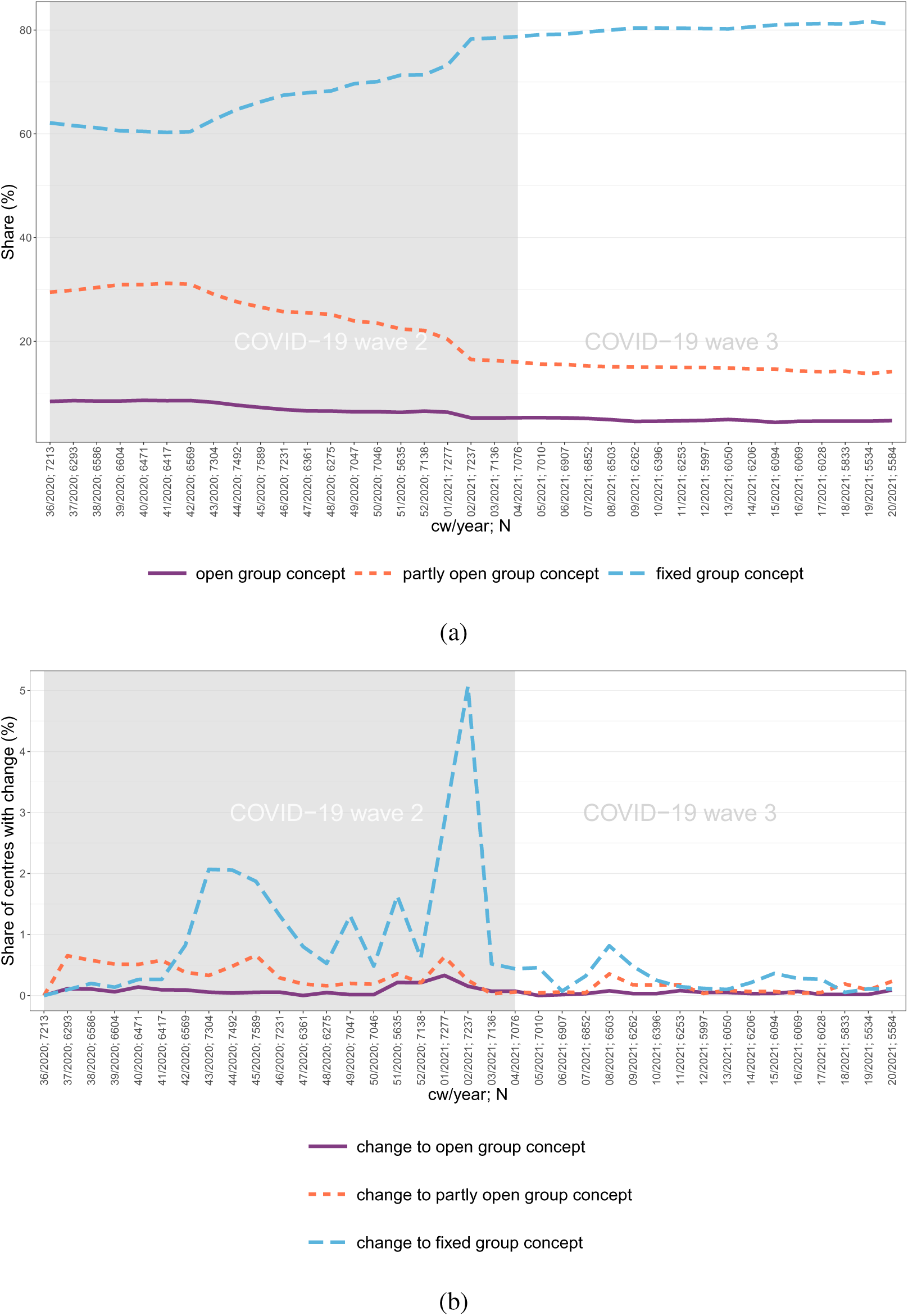
Group concepts: (a) Applied paedagogical group concept in ECEC centres (b) Share of ECEC centres with changes in paedagocial concept (n/N per week) *Source: Survey data collected by the German Youth Institute (ECEC centre registry), own calculations*.

**Figure A5:**
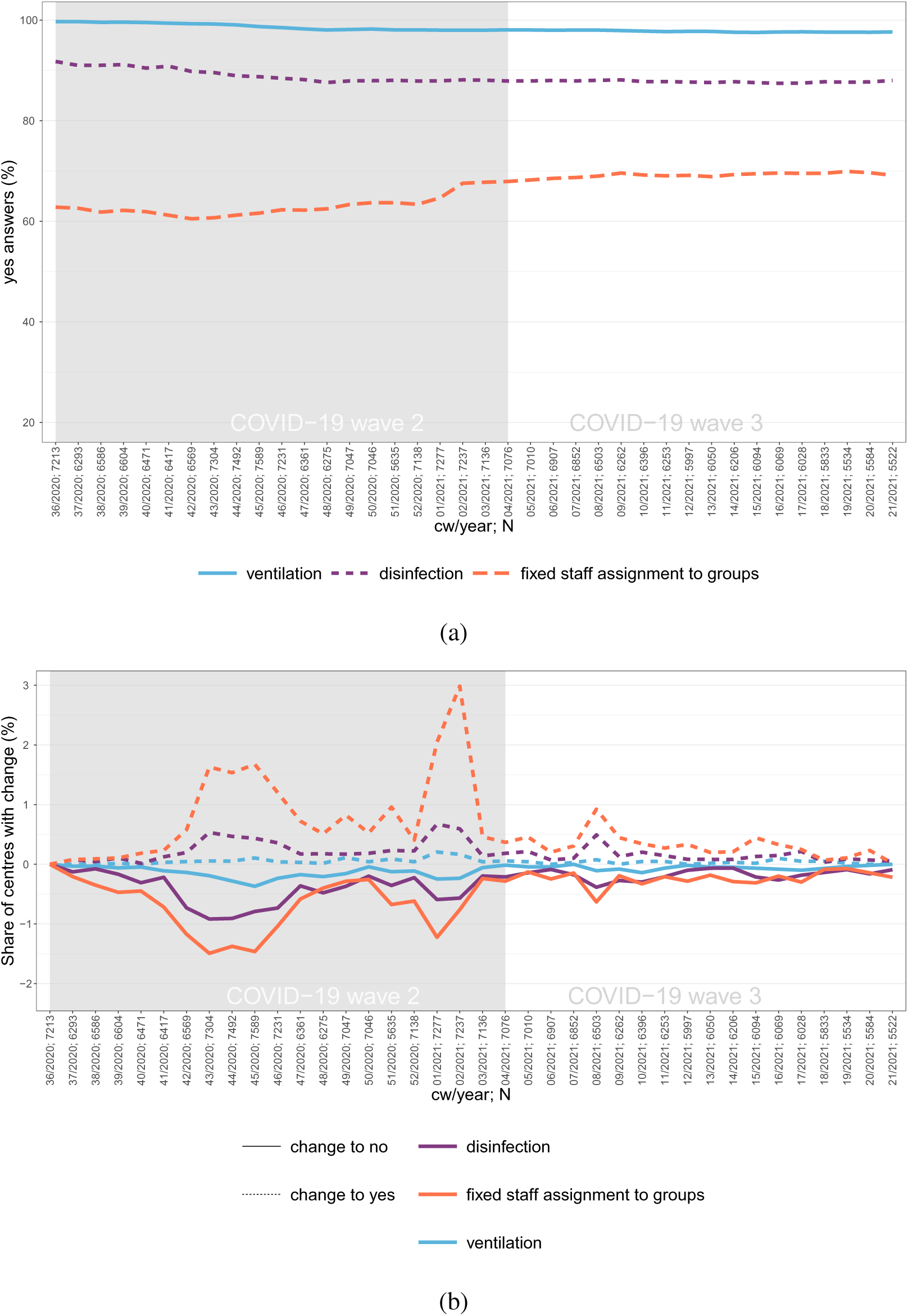
Hygiene measures. (a) Share of ECEC centres with compliance to different hygiene measures (% yes answers) (b) Share of ECEC centres with positive and negative changes in hygiene measures (n/N per week) *Source: Survey data collected by the German Youth Institute (ECEC centre registry), own calculations*.

**Table A2:**
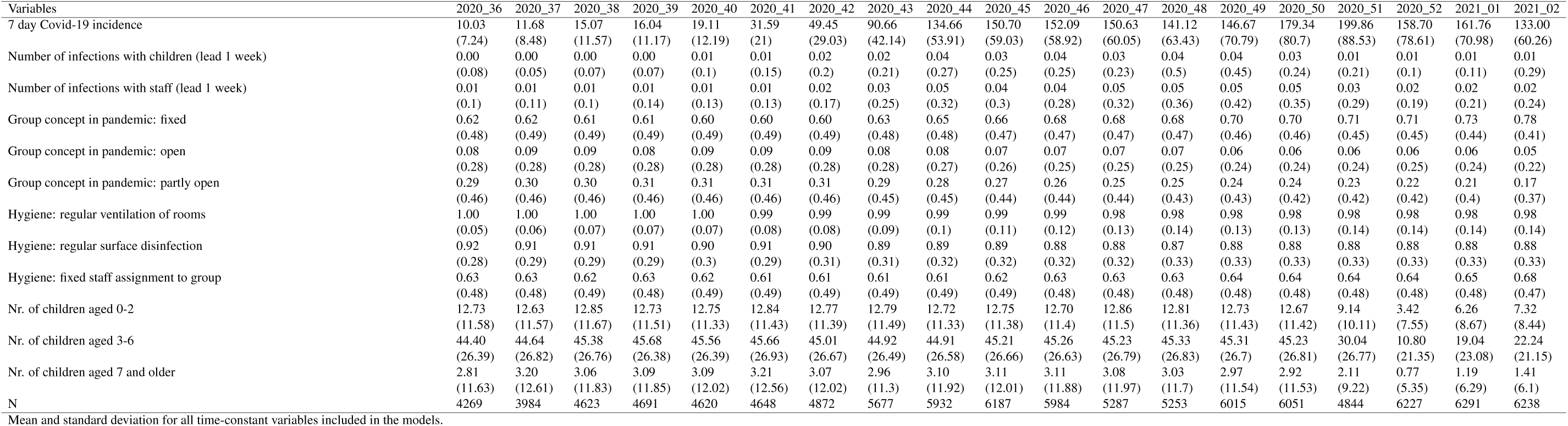
Time-varying variables (part 1, mean, (sd))

**Table A3:**
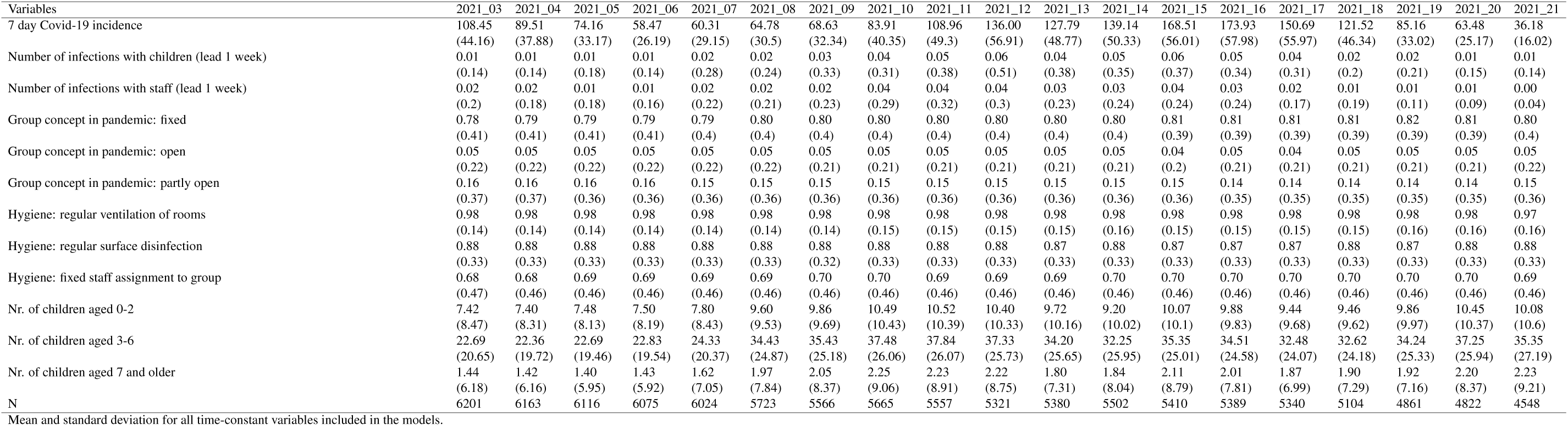
Time-varying variables (part 2, mean, (sd))

**Table A4:** Time-constant variables (mean, (sd))

| Variables | Mean (sd) |
| --- | --- |
| Provider: ecclesiastical | 0.37<br>(0.48) |
| Provider: other non-profit | 0.18<br>(0.38) |
| Provider: private | 0.12<br>(0.33) |
| Provider: public | 0.33<br>(0.47) |
| SES: 0 to 10% of low SES children | 0.52<br>(0.5) |
| SES: 11 to 30% of low SES children | 0.28<br>(0.45) |
| SES: 31 to 60% of low SES children | 0.13<br>(0.34) |
| SES: above 60% of low SES children | 0.08<br>(0.26) |
| Group concept before pandemic: fixed | 0.32<br>(0.47) |
| Group concept before pandemic: open | 0.17<br>(0.37) |
| Group concept before pandemic: partly open | 0.51<br>(0.5) |
| N | 206460 |
| Mean and standard deviation for all time-constant variables included in the models. |  |

**Table A5:**
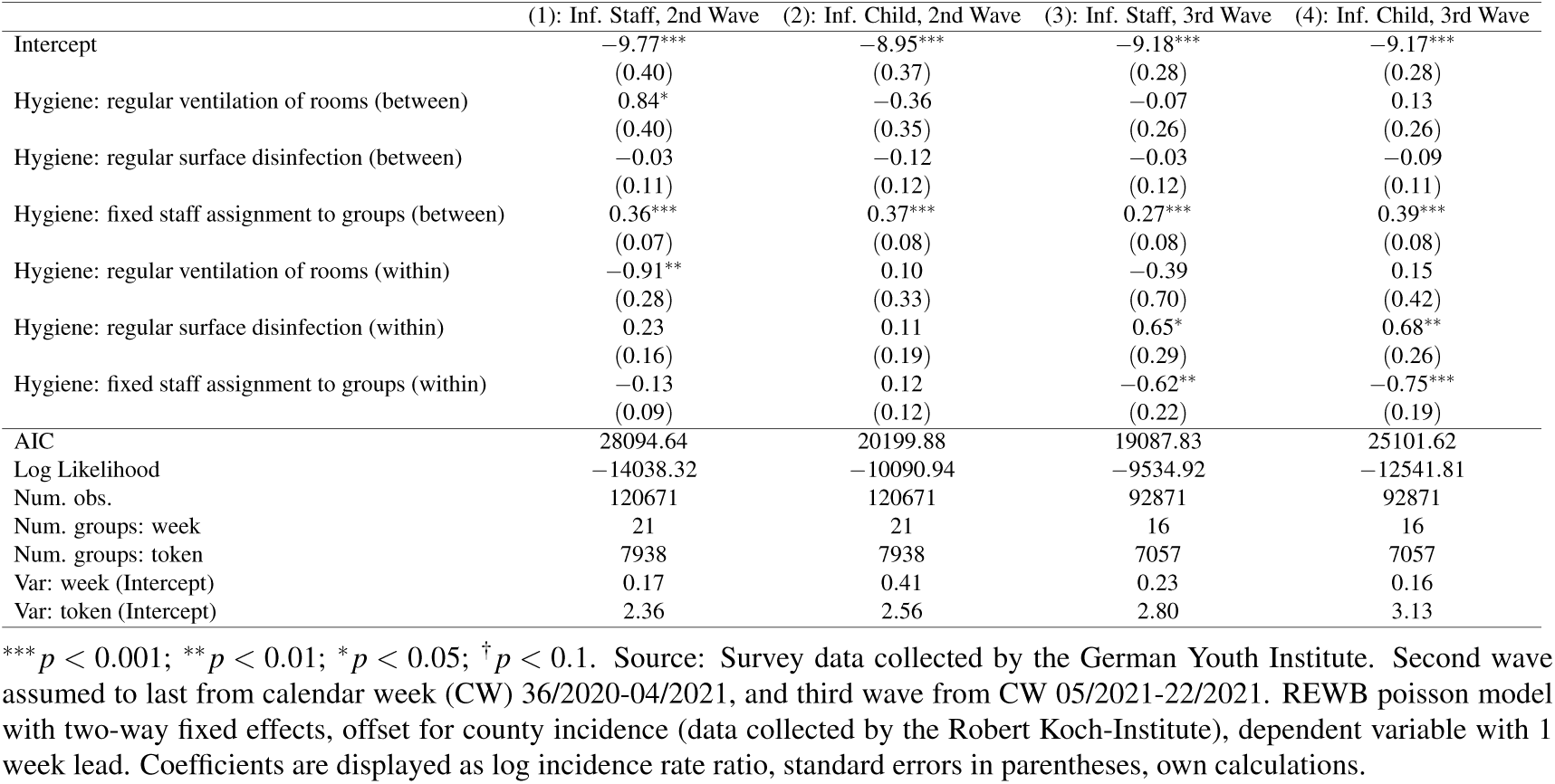
Hygiene variables only: Incidence rate ratios (log) of predictors for infections in staff and children in 2nd (Model 1,2) and 3rd wave (Model 3,4)

|  | (1): Inf. Staff, 2nd Wave | (2): Inf. Child, 2nd Wave | (3): Inf. Staff, 3rd Wave | (4): Inf. Child, 3rd Wave |
| --- | --- | --- | --- | --- |
| Intercept | -9.77***<br>(0.40) | -8.95***<br>(0.37) | -9.18***<br>(0.28) | -9.17***<br>(0.28) |
| Hygiene: regular ventilation of rooms (between) | 0.84*<br>(0.40) | -0.36<br>(0.35) | -0.07<br>(0.26) | 0.13<br>(0.26) |
| Hygiene: regular surface disinfection (between) | -0.03<br>(0.11) | -0.12<br>(0.12) | -0.03<br>(0.12) | -0.09<br>(0.11) |
| Hygiene: fixed staff assignment to groups (between) | 0.36***<br>(0.07) | 0.37***<br>(0.08) | 0.27***<br>(0.08) | 0.39***<br>(0.08) |
| Hygiene: regular ventilation of rooms (within) | -0.91**<br>(0.28) | 0.10<br>(0.33) | -0.39<br>(0.70) | 0.15<br>(0.42) |
| Hygiene: regular surface disinfection (within) | 0.23<br>(0.16) | 0.11<br>(0.19) | 0.65*<br>(0.29) | 0.68**<br>(0.26) |
| Hygiene: fixed staff assignment to groups (within) | -0.13<br>(0.09) | 0.12<br>(0.12) | -0.62**<br>(0.22) | -0.75***<br>(0.19) |
| AIC | 28094.64 | 20199.88 | 19087.83 | 25101.62 |
| Log Likelihood | -14038.32 | -10090.94 | -9534.92 | -12541.81 |
| Num. obs. | 120671 | 120671 | 92871 | 92871 |
| Num. groups: week | 21 | 21 | 16 | 16 |
| Num. groups: token | 7938 | 7938 | 7057 | 7057 |
| Var: week (Intercept) | 0.17 | 0.41 | 0.23 | 0.16 |
| Var: token (Intercept) | 2.36 | 2.56 | 2.80 | 3.13 |

\*\*\* $p < 0.001$ ; \*\* $p < 0.01$ ; \* $p < 0.05$ ; $^{\dagger} p < 0.1$ . Source: Survey data collected by the German Youth Institute. Second wave assumed to last from calendar week (CW) 36/2020-04/2021, and third wave from CW 05/2021-22/2021. REWB poisson model with two-way fixed effects, offset for county incidence (data collected by the Robert Koch-Institute), dependent variable with 1 week lead. Coefficients are displayed as log incidence rate ratio, standard errors in parentheses, own calculations.

**Table A6:** Ventilation only: Incidence rate ratios (log) of predictors for infections in staff and children in 2nd (Model 1,2) and 3rd wave (Model 3,4)

|  | (1): Inf. Staff, 2nd Wave | (2): Inf. Child, 2nd Wave | (3): Inf. Staff, 3rd Wave | (4): Inf. Child, 3rd Wave |
| --- | --- | --- | --- | --- |
| Intercept | -9.61***<br>(0.40) | -8.81***<br>(0.37) | -9.01***<br>(0.27) | -8.94***<br>(0.27) |
| Hygiene: regular ventilation of rooms (between) | 0.89*<br>(0.39) | -0.37<br>(0.34) | -0.06<br>(0.25) | 0.10<br>(0.25) |
| Hygiene: regular ventilation of rooms (within) | -0.78**<br>(0.26) | 0.18<br>(0.30) | -0.22<br>(0.68) | 0.22<br>(0.42) |
| AIC | 28389.33 | 20394.06 | 19388.76 | 25538.28 |
| Log Likelihood | -14189.67 | -10192.03 | -9689.38 | -12764.14 |
| Num. obs. | 121704 | 121704 | 94412 | 94412 |
| Num. groups: week | 21 | 21 | 16 | 16 |
| Num. groups: token | 7942 | 7942 | 7157 | 7157 |
| Var: week (Intercept) | 0.17 | 0.41 | 0.23 | 0.16 |
| Var: token (Intercept) | 2.38 | 2.57 | 2.81 | 3.14 |

\*\*\* $p < 0.001$ ; \*\* $p < 0.01$ ; \* $p < 0.05$ ; $^{\dagger} p < 0.1$ . Source: Survey data collected by the German Youth Institute. Second wave assumed to last from calendar week (CW) 36/2020-04/2021, and third wave from CW 05/2021-22/2021. REWB poisson model with two-way fixed effects, offset for county incidence (data collected by the Robert Koch-Institute), dependent variable with 1 week lead. Coefficients are displayed as log incidence rate ratio, standard errors in parentheses, own calculations.

**Table A7:** Desinfection only: Incidence rate ratios (log) of predictors for infections in staff and children in 2nd (Model 1,2) and 3rd wave (Model 3,4)

|  | (1): Inf. Staff, 2nd Wave | (2): Inf. Child, 2nd Wave | (3): Inf. Staff, 3rd Wave | (4): Inf. Child, 3rd Wave |
| --- | --- | --- | --- | --- |
| Intercept | −8.73***<br>(0.13) | −9.06***<br>(0.18) | −9.05***<br>(0.16) | −8.78***<br>(0.14) |
| Hygiene: regular surface disinfection (between) | −0.00<br>(0.10) | −0.13<br>(0.12) | −0.03<br>(0.11) | −0.07<br>(0.11) |
| Hygiene: regular surface disinfection (within) | 0.07<br>(0.15) | 0.14<br>(0.18) | 0.56*<br>(0.28) | 0.64*<br>(0.25) |
| AIC | 28406.65 | 20396.73 | 19388.15 | 25534.27 |
| Log Likelihood | −14198.33 | −10193.37 | −9689.08 | −12762.13 |
| Num. obs. | 121704 | 121704 | 94412 | 94412 |
| Num. groups: week | 21 | 21 | 16 | 16 |
| Num. groups: token | 7942 | 7942 | 7157 | 7157 |
| Var: week (Intercept) | 0.17 | 0.41 | 0.23 | 0.16 |
| Var: token (Intercept) | 2.39 | 2.57 | 2.81 | 3.14 |

\*\*\* $p < 0.001$ ; \*\* $p < 0.01$ ; \* $p < 0.05$ ; † $p < 0.1$ . Source: Survey data collected by the German Youth Institute. Second wave assumed to last from calendar week (CW) 36/2020-04/2021, and third wave from CW 05/2021-22/2021. REWB poisson model with two-way fixed effects, offset for county incidence (data collected by the Robert Koch-Institute), dependent variable with 1 week lead. Coefficients are displayed as log incidence rate ratio, standard errors in parentheses, own calculations.

**Table A8:** Staff assignment only: Incidence rate ratios (log) of predictors for infections in staff and children in 2nd (Model 1,2) and 3rd wave (Model 3,4)

|  | (1): Inf. Staff, 2nd Wave | (2): Inf. Child, 2nd Wave | (3): Inf. Staff, 3rd Wave | (4): Inf. Child, 3rd Wave |
| --- | --- | --- | --- | --- |
| Intercept | −8.97***<br>(0.11) | −9.41***<br>(0.16) | −9.27***<br>(0.14) | −9.13***<br>(0.12) |
| Hygiene: fixed staff assignment to groups (between) | 0.37***<br>(0.07) | 0.37***<br>(0.08) | 0.28***<br>(0.08) | 0.40***<br>(0.08) |
| Hygiene: fixed staff assignment to groups (within) | −0.13<br>(0.09) | 0.12<br>(0.12) | −0.59**<br>(0.22) | −0.71***<br>(0.19) |
| AIC | 28113.28 | 20207.05 | 19085.70 | 25101.69 |
| Log Likelihood | −14051.64 | −10098.52 | −9537.85 | −12545.85 |
| Num. obs. | 120987 | 120987 | 93030 | 93030 |
| Num. groups: week | 21 | 21 | 16 | 16 |
| Num. groups: token | 7953 | 7953 | 7070 | 7070 |
| Var: week (Intercept) | 0.17 | 0.41 | 0.23 | 0.16 |
| Var: token (Intercept) | 2.37 | 2.56 | 2.80 | 3.12 |

\*\*\* $p < 0.001$ ; \*\* $p < 0.01$ ; \* $p < 0.05$ ; † $p < 0.1$ . Source: Survey data collected by the German Youth Institute (ECEC centre registry). Second wave assumed to last from calendar week (CW) 36/2020-04/2021, and third wave from CW 05/2021-22/2021. REWB poisson model with two-way fixed effects, offset for county incidence (data collected by the Robert Koch-Institute), dependent variable with 1 week lead. Coefficients are displayed as log incidence rate ratio, standard errors in parentheses, own calculations.

**Table A9:**
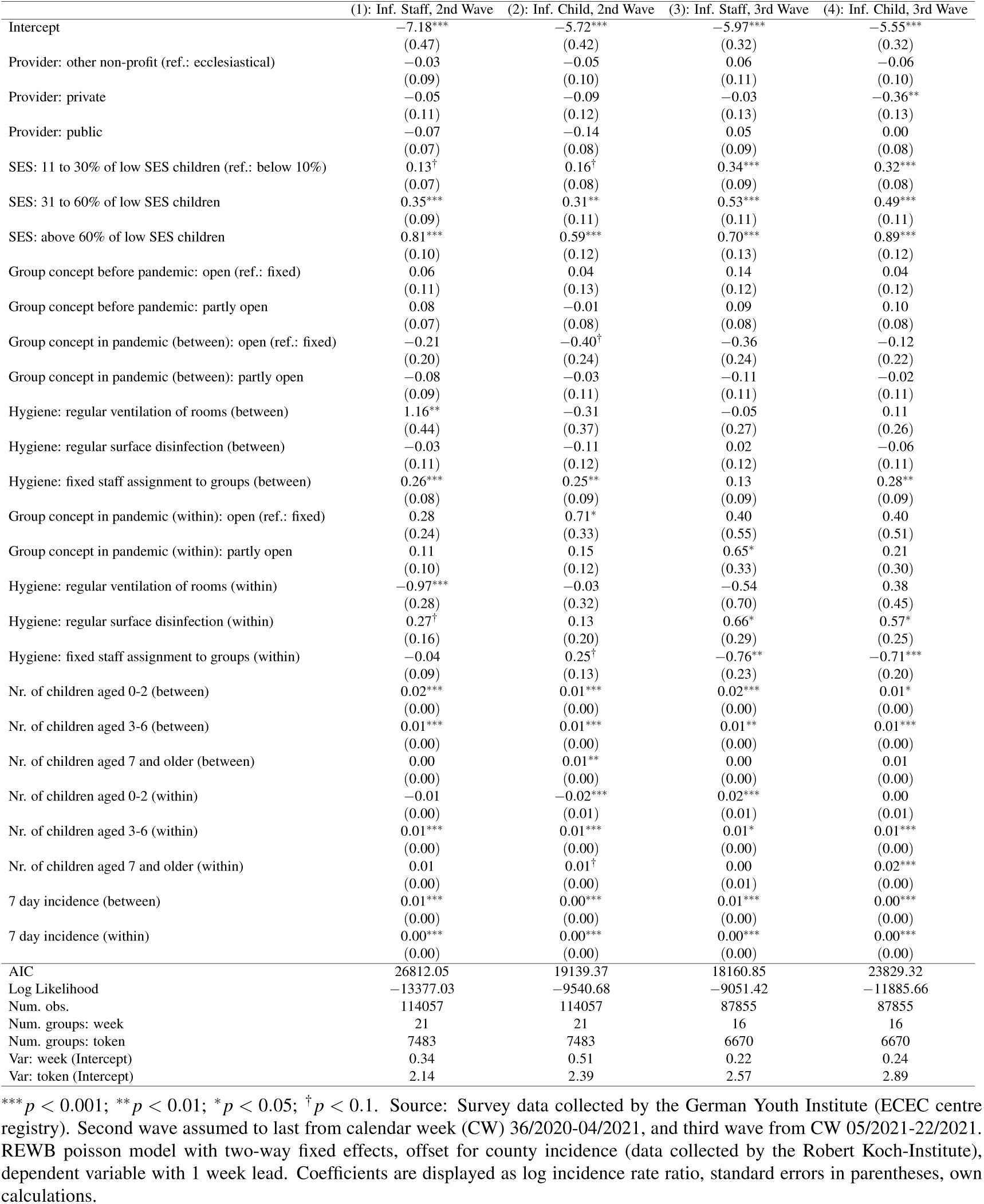
Offset (7 day incidence) as variable: Incidence rate ratios (log) of predictors for infections in staff and children in 2nd (Model 1,2) and 3rd wave (Model 3,4)

**Table A10:**
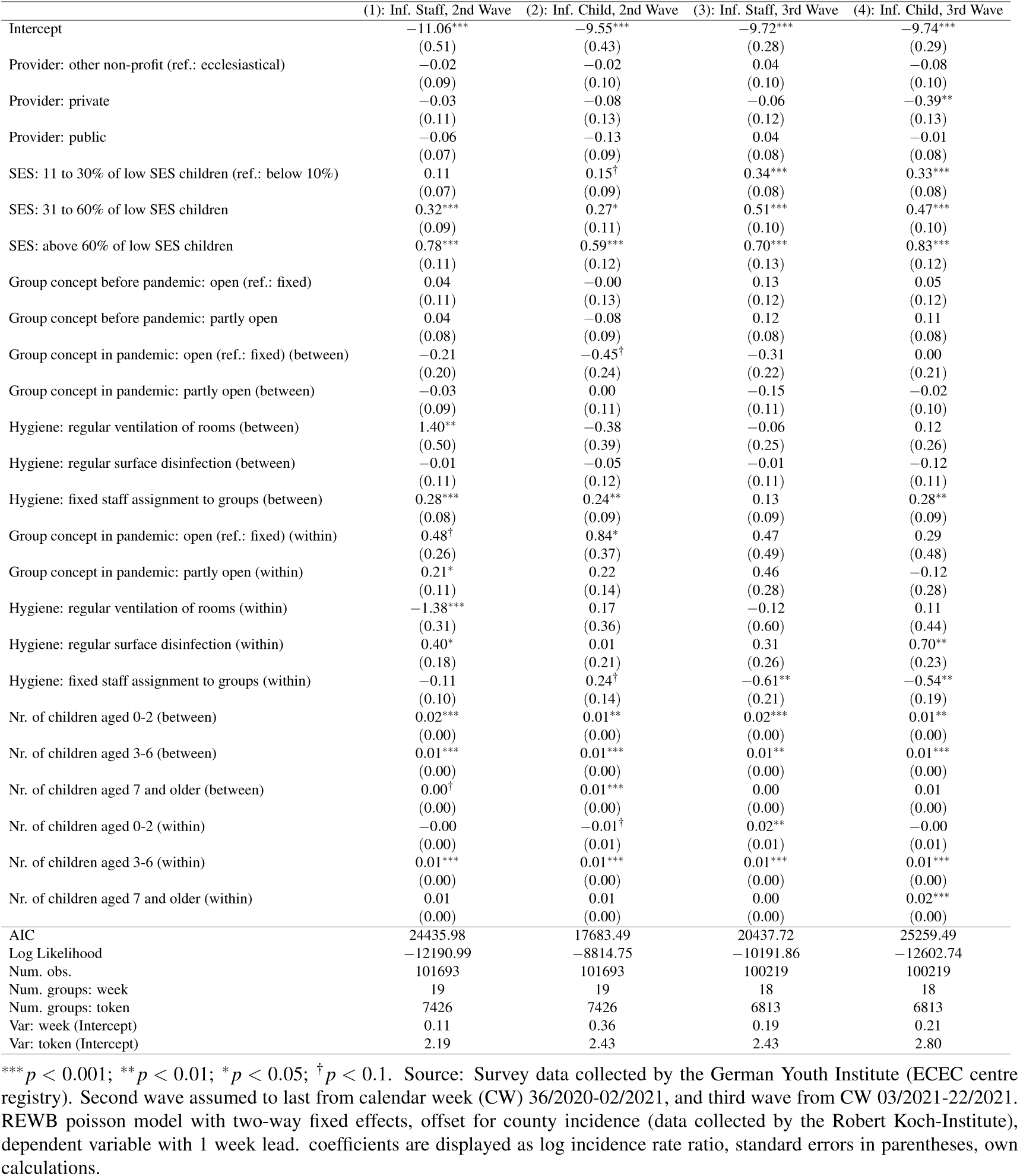
Alternative Wave Definition: Incidence rate ratios (log) of predictors for infections in staff and children in 2nd (Model 1,2) and 3rd wave (Model 3,4)

**Table A11:** Alternative Wave Definition 2: Incidence rate ratios (log) of predictors for infections in staff and children in 2nd (Model 1,2) and 3rd wave (Model 3,4)

|  | (1): Inf. Staff, 2nd Wave | (2): Inf. Child, 2nd Wave | (3): Inf. Staff, 3rd Wave | (4): Inf. Child, 3rd Wave |
| --- | --- | --- | --- | --- |
| Intercept | -10.75***<br>(0.42) | -9.56***<br>(0.38) | -9.78***<br>(0.31) | -9.66***<br>(0.29) |
| Provider: other non-profit (ref.: ecclesiastical) | -0.04<br>(0.08) | -0.04<br>(0.10) | 0.08<br>(0.11) | -0.06<br>(0.11) |
| Provider: private | -0.07<br>(0.10) | -0.14<br>(0.12) | -0.01<br>(0.14) | -0.35*<br>(0.14) |
| Provider: public | -0.07<br>(0.07) | -0.11<br>(0.08) | 0.07<br>(0.09) | 0.01<br>(0.08) |
| SES: 11 to 30% of low SES children (ref.: below 10%) | 0.14*<br>(0.07) | 0.17*<br>(0.08) | 0.34***<br>(0.09) | 0.32***<br>(0.09) |
| SES: 31 to 60% of low SES children | 0.37***<br>(0.09) | 0.31**<br>(0.10) | 0.51***<br>(0.11) | 0.47***<br>(0.11) |
| SES: above 60% of low SES children | 0.76***<br>(0.10) | 0.56***<br>(0.12) | 0.73***<br>(0.14) | 0.87***<br>(0.13) |
| Group concept before pandemic: open (ref.: fixed) | 0.03<br>(0.11) | 0.03<br>(0.12) | 0.14<br>(0.13) | 0.02<br>(0.12) |
| Group concept before pandemic: partly open | 0.07<br>(0.07) | -0.02<br>(0.08) | 0.05<br>(0.09) | 0.06<br>(0.08) |
| Group concept in pandemic: open (ref.: fixed) (between) | -0.19<br>(0.20) | -0.45†<br>(0.24) | -0.43†<br>(0.25) | -0.06<br>(0.23) |
| Group concept in pandemic: partly open (between) | -0.09<br>(0.09) | -0.05<br>(0.11) | -0.07<br>(0.11) | 0.01<br>(0.11) |
| Hygiene: regular ventilation of rooms (between) | 1.10**<br>(0.41) | -0.34<br>(0.35) | -0.08<br>(0.27) | 0.16<br>(0.27) |
| Hygiene: regular surface disinfection (between) | 0.01<br>(0.10) | -0.11<br>(0.12) | -0.00<br>(0.13) | -0.08<br>(0.12) |
| Hygiene: fixed staff assignment to groups (between) | 0.23**<br>(0.08) | 0.24**<br>(0.09) | 0.12<br>(0.09) | 0.26**<br>(0.09) |
| Group concept in pandemic: open (ref.: fixed) (within) | 0.25<br>(0.23) | 0.63*<br>(0.31) | 0.53<br>(0.62) | 0.80<br>(0.56) |
| Group concept in pandemic: partly open (within) | 0.15<br>(0.09) | 0.25*<br>(0.12) | 0.38<br>(0.39) | 0.32<br>(0.35) |
| Hygiene: regular ventilation of rooms (within) | -0.79**<br>(0.27) | -0.05<br>(0.31) | -0.62<br>(0.77) | 0.43<br>(0.48) |
| Hygiene: regular surface disinfection (within) | 0.23<br>(0.16) | 0.20<br>(0.19) | 0.75*<br>(0.32) | 0.74**<br>(0.28) |
| Hygiene: fixed staff assignment to groups (within) | -0.05<br>(0.09) | 0.34**<br>(0.12) | -0.65*<br>(0.26) | -0.69**<br>(0.22) |
| Nr. of children aged 0-2 (between) | 0.03***<br>(0.00) | 0.01***<br>(0.00) | 0.02***<br>(0.00) | 0.01*<br>(0.00) |
| Nr. of children aged 3-6 (between) | 0.01***<br>(0.00) | 0.01***<br>(0.00) | 0.01***<br>(0.00) | 0.01***<br>(0.00) |
| Nr. of children aged 7 and older (between) | 0.01†<br>(0.00) | 0.01**<br>(0.00) | 0.00<br>(0.00) | 0.01<br>(0.00) |
| Nr. of children aged 0-2 (within) | -0.00<br>(0.00) | -0.02***<br>(0.01) | 0.01<br>(0.01) | -0.00<br>(0.01) |
| Nr. of children aged 3-6 (within) | 0.01***<br>(0.00) | 0.01***<br>(0.00) | 0.00†<br>(0.00) | 0.01***<br>(0.00) |
| Nr. of children aged 7 and older (within) | 0.00<br>(0.00) | 0.01***<br>(0.00) | 0.01<br>(0.01) | 0.02***<br>(0.00) |
| AIC | 28595.74 | 20601.74 | 16384.26 | 22294.80 |
| Log Likelihood | -14270.87 | -10273.87 | -8165.13 | -11120.40 |
| Num. obs. | 126248 | 126248 | 75664 | 75664 |
| Num. groups: week | 23 | 23 | 14 | 14 |
| Num. groups: token | 7535 | 7535 | 6543 | 6543 |
| Var: week (Intercept) | 0.10 | 0.28 | 0.24 | 0.09 |
| Var: token (Intercept) | 2.03 | 2.34 | 2.68 | 3.00 |

\*\*\* $p < 0.001$ ; \*\* $p < 0.01$ ; \* $p < 0.05$ ; † $p < 0.1$ . Source: Survey data collected by the German Youth Institute (ECEC centre registry). Second wave assumed to last from calendar week (CW) 36/2020-06/2021, and third wave from CW 07/2021-22/2021. REWB poisson model with two-way fixed effects, offset for county incidence (data collected by the Robert Koch-Institute), dependent variable with 1 week lead. coefficients are displayed as log incidence rate ratio, standard errors in parentheses, own calculations.

**Table A12:**
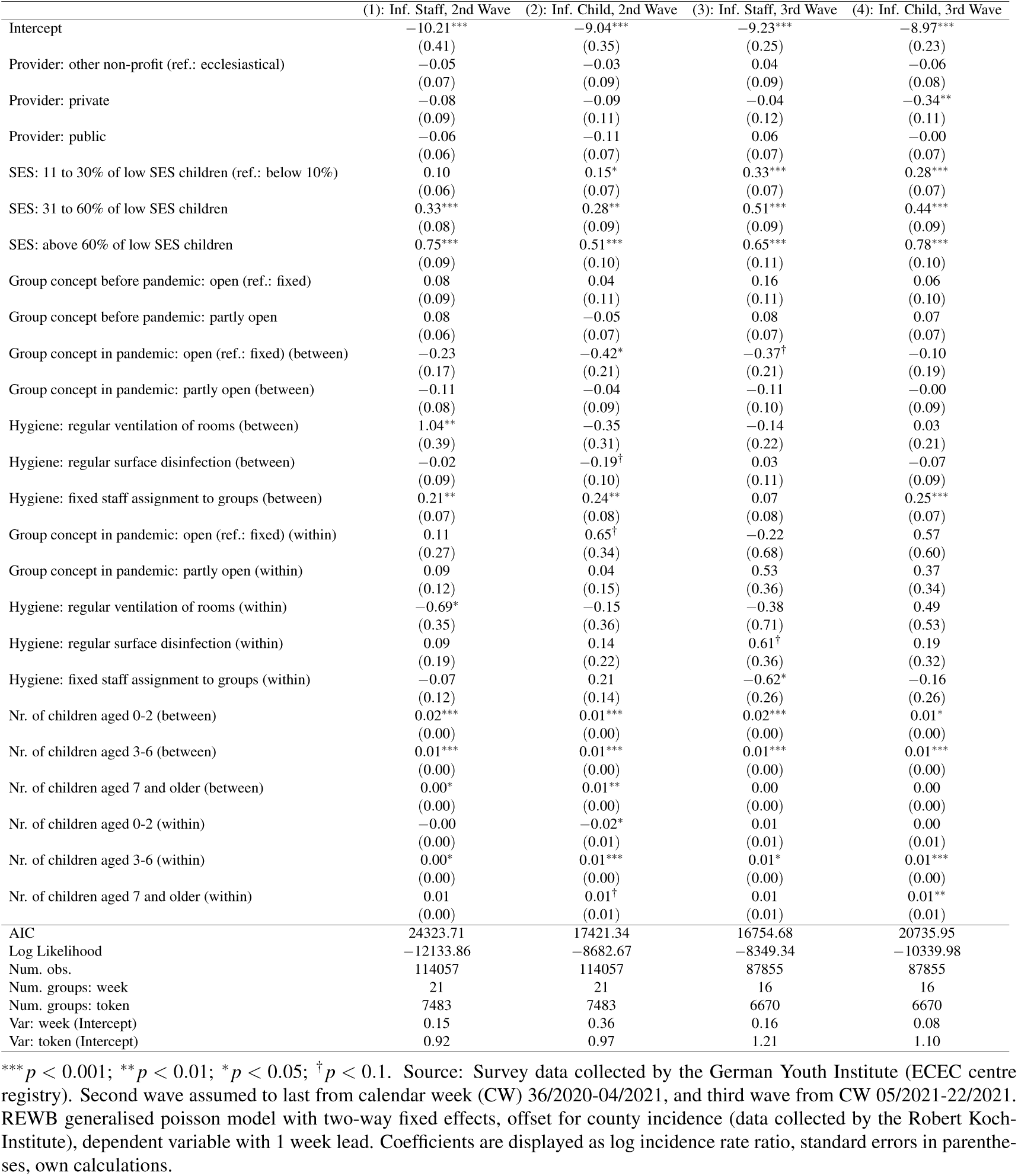
Alternative Distribution (Generalised Poisson): Incidence rate ratios (log) of predictors for infections in staff and children in 2nd (Model 1,2) and 3rd wave (Model 3,4)

|  | (1): Inf. Staff, 2nd Wave | (2): Inf. Child, 2nd Wave | (3): Inf. Staff, 3rd Wave | (4): Inf. Child, 3rd Wave |
| --- | --- | --- | --- | --- |
| Intercept | -10.21***<br>(0.41) | -9.04***<br>(0.35) | -9.23***<br>(0.25) | -8.97***<br>(0.23) |
| Provider: other non-profit (ref.: ecclesiastical) | -0.05<br>(0.07) | -0.03<br>(0.09) | 0.04<br>(0.09) | -0.06<br>(0.08) |
| Provider: private | -0.08<br>(0.09) | -0.09<br>(0.11) | -0.04<br>(0.12) | -0.34**<br>(0.11) |
| Provider: public | -0.06<br>(0.06) | -0.11<br>(0.07) | 0.06<br>(0.07) | -0.00<br>(0.07) |
| SES: 11 to 30% of low SES children (ref.: below 10%) | 0.10<br>(0.06) | 0.15*<br>(0.07) | 0.33***<br>(0.07) | 0.28***<br>(0.07) |
| SES: 31 to 60% of low SES children | 0.33***<br>(0.08) | 0.28**<br>(0.09) | 0.51***<br>(0.09) | 0.44***<br>(0.09) |
| SES: above 60% of low SES children | 0.75***<br>(0.09) | 0.51***<br>(0.10) | 0.65***<br>(0.11) | 0.78***<br>(0.10) |
| Group concept before pandemic: open (ref.: fixed) | 0.08<br>(0.09) | 0.04<br>(0.11) | 0.16<br>(0.11) | 0.06<br>(0.10) |
| Group concept before pandemic: partly open | 0.08<br>(0.06) | -0.05<br>(0.07) | 0.08<br>(0.07) | 0.07<br>(0.07) |
| Group concept in pandemic: open (ref.: fixed) (between) | -0.23<br>(0.17) | -0.42*<br>(0.21) | -0.37†<br>(0.21) | -0.10<br>(0.19) |
| Group concept in pandemic: partly open (between) | -0.11<br>(0.08) | -0.04<br>(0.09) | -0.11<br>(0.10) | -0.00<br>(0.09) |
| Hygiene: regular ventilation of rooms (between) | 1.04**<br>(0.39) | -0.35<br>(0.31) | -0.14<br>(0.22) | 0.03<br>(0.21) |
| Hygiene: regular surface disinfection (between) | -0.02<br>(0.09) | -0.19†<br>(0.10) | 0.03<br>(0.11) | -0.07<br>(0.09) |
| Hygiene: fixed staff assignment to groups (between) | 0.21**<br>(0.07) | 0.24**<br>(0.08) | 0.07<br>(0.08) | 0.25***<br>(0.07) |
| Group concept in pandemic: open (ref.: fixed) (within) | 0.11<br>(0.27) | 0.65†<br>(0.34) | -0.22<br>(0.68) | 0.57<br>(0.60) |
| Group concept in pandemic: partly open (within) | 0.09<br>(0.12) | 0.04<br>(0.15) | 0.53<br>(0.36) | 0.37<br>(0.34) |
| Hygiene: regular ventilation of rooms (within) | -0.69*<br>(0.35) | -0.15<br>(0.36) | -0.38<br>(0.71) | 0.49<br>(0.53) |
| Hygiene: regular surface disinfection (within) | 0.09<br>(0.19) | 0.14<br>(0.22) | 0.61†<br>(0.36) | 0.19<br>(0.32) |
| Hygiene: fixed staff assignment to groups (within) | -0.07<br>(0.12) | 0.21<br>(0.14) | -0.62*<br>(0.26) | -0.16<br>(0.26) |
| Nr. of children aged 0-2 (between) | 0.02***<br>(0.00) | 0.01***<br>(0.00) | 0.02***<br>(0.00) | 0.01*<br>(0.00) |
| Nr. of children aged 3-6 (between) | 0.01***<br>(0.00) | 0.01***<br>(0.00) | 0.01***<br>(0.00) | 0.01***<br>(0.00) |
| Nr. of children aged 7 and older (between) | 0.00*<br>(0.00) | 0.01**<br>(0.00) | 0.00<br>(0.00) | 0.00<br>(0.00) |
| Nr. of children aged 0-2 (within) | -0.00<br>(0.00) | -0.02*<br>(0.01) | 0.01<br>(0.01) | 0.00<br>(0.01) |
| Nr. of children aged 3-6 (within) | 0.00*<br>(0.00) | 0.01***<br>(0.00) | 0.01*<br>(0.00) | 0.01***<br>(0.00) |
| Nr. of children aged 7 and older (within) | 0.01<br>(0.00) | 0.01†<br>(0.01) | 0.01<br>(0.01) | 0.01**<br>(0.01) |
| AIC | 24323.71 | 17421.34 | 16754.68 | 20735.95 |
| Log Likelihood | -12133.86 | -8682.67 | -8349.34 | -10339.98 |
| Num. obs. | 114057 | 114057 | 87855 | 87855 |
| Num. groups: week | 21 | 21 | 16 | 16 |
| Num. groups: token | 7483 | 7483 | 6670 | 6670 |
| Var: week (Intercept) | 0.15 | 0.36 | 0.16 | 0.08 |
| Var: token (Intercept) | 0.92 | 0.97 | 1.21 | 1.10 |

\*\*\* $p < 0.001$ ; \*\* $p < 0.01$ ; \* $p < 0.05$ ; † $p < 0.1$ . Source: Survey data collected by the German Youth Institute (ECEC centre registry). Second wave assumed to last from calendar week (CW) 36/2020-04/2021, and third wave from CW 05/2021-22/2021. REWB generalised poisson model with two-way fixed effects, offset for county incidence (data collected by the Robert Koch-Institute), dependent variable with 1 week lead. Coefficients are displayed as log incidence rate ratio, standard errors in parentheses, own calculations.

## Footnotes

1 https://www.gesetze-im-internet.de/ifsg/, visited 13.4.2021

## Notes

### Competing Interest Statement

The authors have declared no competing interest.

### Funding Statement

The research presented in this work was supported by the German Federal Ministry of Family Affairs (Bundesministeriums fuer Familie, Senioren, Frauen und Jugend) and the German Federal Ministry of Health (Bundesministerium fuer Gesundheit).

### Author Declarations

Ethical approval for the study was waived by the Ethics Committee of the German Youth Institute (Head: Sarah Hanke; Members: Christina Boll, PhD; Kathja Flaeming, PhD; Peter Furthmueller, PhD; Heinz Kindler, Prof.; Jens Pothmann, PhD; Frank Tillmann, PhD; Sabine Walter, Prof.; Herwig Reiter, PhD; all German Youth Institute). We do not require a vote of the ethics committee as the data used in this study is survey data from organizations only (day care centers) and in no way assignable to persons. The only ethically relevant points are questions of privacy and data protection. All data is anonymised and subject to strict data protection guidelines, as approved by the data protection experts of the German Youth Institute, the German Federal Ministry of Family Affairs (Bundesministerium fuer Familie, Senioren, Frauen und Jugend) and the Federal Commissioner for Data Protection and Freedom of Information (Bundesbeautragter fuer den Datenschutz und die Informationsfreiheit).

### Summary of Updates

The underlying data now cover the entire 3rd wave of the pamdemic. In addition, the presentation of the results has been improved.

## References

[1] Autorengruppe Corona-KiTa-Studie. Monatsbericht der Corona-KiTa-Studie (Vol. 1/2020 - 10/2020). Deutsches Jugendinstitut und Robert Koch-Institut; 2020. Available from: https://corona-kita-studie.de/ergebnisse#berichte.

[2] Autorengruppe Corona-KiTa-Studie. Quartalsbericht der Corona-KiTa-Studie (Vol. III/2020 - IV/2020). Deutsches Jugendinstitut und Robert Koch-Institut; 2020. Available from: https://corona-kita-studie.de/ergebnisse#berichte.

[3] Autorengruppe Corona-KiTa-Studie. Monatsbericht 1/2021 der Corona-KiTa-Studie. Deutsches Jugendinstitut und Robert Koch-Institut; 2021. Available from: https://corona-kita-studie.de/ergebnisse#berichte.

[4] Autorengruppe Corona-KiTa-Studie. Quartalsbericht I/2021 der Corona-KiTa-Studie. Deutsches Jugendinstitut und Robert Koch-Institut; 2021. Available from: https://corona-kita-studie.de/ergebnisse#berichte.

[5] JFMK. Beschluss der Jugend- und Familienministerkonferenz (JFMK) gemeinsam mit der Bundesministerin für Familie, Senioren, Frauen und Jugend vom 28.04.2020; 2020.

[6] European Centre for Disease Prevention and Control. COVID-19 in children (1-18 years) and the role of school settings in COVID-19 transmission : 1 st update. European Centre for Disease Prevention and Control; 2020.

[7] Viner RM, Mytton OT, Bonell C, Melendez-Torres GJ, Ward J, Hudson L, et al. Susceptibility to SARS-CoV-2 Infection among Children and Adolescents Compared with Adults: A Systematic Review and Meta-Analysis. JAMA Pediatrics. 2021;175(2):143–156.

[8] Koh WC, Naing L, Chaw L, Rosledzana MA, Alikhan MF, Jamaludin SA, et al. What do we know about SARS-CoV-2 transmission? A systematic review and meta-analysis of the secondary attack rate and associated risk factors. PLoS ONE. 2020;15(10):1–23. Available from: http://dx.doi.org/10.1371/journal.pone.0240205.

[9] Madewell ZJ, Yang Y, Longini IM, Halloran ME, Dean NE. Household Transmission of SARS-CoV-2: A Systematic Review and Meta-analysis. JAMA network open. 2020;3(12):e2031756.

[10] Schoeps A, Hoffmann D, Tamm C, Vollmer B, Haag S, Kaffenberger T, et al. COVID-19 Transmission in Educational Institutions August to December 2020, Rhineland-Palatinate, Germany: A Study of Index Cases and Close Contact Cohorts. SSRN Electronic Journal. 2021;(December 2020).

[11] Robert Koch-Institut. Epidemiologischer Steckbrief zu SARS-CoV-2 und COVID-19; 2021. Available from: https://www.rki.de/DE/Content/InfAZ/N/Neuartiges_Coronavirus/Steckbrief.html.

[12] Azimi P, Keshavarz Z, Cedeno Laurent JG, Stephens B, Allen JG. Mechanistic transmission modeling of COVID-19 on the Diamond Princess cruise ship demonstrates the importance of aerosol transmission. Proceedings of the National Academy of Sciences. 2021 feb;118(8):e2015482118. Available from: http://www.pnas.org/content/118/8/e2015482118.abstract.

[13] Meyerowitz EA, Richterman A, Gandhi RT, Sax PE. Transmission of SARS-CoV-2: A Review of Viral, Host, and Environmental Factors. Annals of internal medicine. 2021 jan;174(1):69–79.

[14] Buonanno G, Stabile L, Morawska L. Estimation of airborne viral emission: Quanta emission rate of SARS-CoV-2 for infection risk assessment. Environment International. 2020;141(May):105794. Available from: https://doi.org/10.1016/j.envint.2020.105794.

[15] van Doremalen N, Bushmaker T, Morris DH, Holbrook MG, Gamble A, Williamson BN, et al. Aerosol and Surface Stability of SARS-CoV-2 as Compared with SARS-CoV-1. Nejm. 2020;p. 0–2.

[16] Kriegel M, Buchholz U, Gastmeier P, Bischoff P, Abdelgawad I, Hartmann A. Predicted infection risk for aerosol transmission of sars-COV-2. medRxiv. 2020;.

[17] Ferretti L, Wymant C, Kendall M, Zhao L, Nurtay A, Abeler-Dörner L, et al. Quantifying SARS-CoV-2 transmission suggests epidemic control with digital contact tracing. medRxiv. 2020;6936(March):1–13.

[18] Wang Y, Tian H, Zhang L, Zhang M, Guo D, Wu W, et al. Reduction of secondary transmission of SARS-CoV-2 in households by face mask use, disinfection and social distancing: a cohort study in Beijing, China. BMJ Global Health. 2020;5(5):1–9.

[19] BMFSFJ; BMG. Kitas in Zeiten der Corona-Pandemie. Praxistipps für die Kindertagesbetreuung im Regelbetrieb. BMFSFJ; BMG; 2020.

[20] Robert Koch-Institut. Bericht zu Virusvarianten von SARS-CoV-2 in Deutschland, insbesondere zur Variant of Concern (VOC) B.1.1.7. Robert Koch-Institut; 2021. i.

[21] Rai B, Shukla A, Dwivedi LK. Estimates of serial interval for COVID-19: A systematic review and meta-analysis. Clinical Epidemiology and Global Health. 2021;9(January):157–161.

[22] Alene M, Yismaw L, Assemie MA, Ketema DB, Gietaneh W, Birhan TY. Serial interval and incubation period of COVID-19: a systematic review and meta-analysis. BMC Infectious Diseases. 2021;21(1):1–9.

[23] Böhmer MM, Buchholz U, Corman VM, Hoch M, Katz K, Marosevic DV, et al. Investigation of a COVID-19 outbreak in Germany resulting from a single travel-associated primary case: a case series. The Lancet Infectious Diseases. 2020;20(8):920–928.

[24] Baena-Diéz JM, Barroso M, Cordeiro-Coelho SI, Diáz JL, Grau M. Impact of COVID-19 outbreak by income: Hitting hardest the most deprived. Journal of Public Health (United Kingdom). 2020;42(4):698–703.

[25] Wachtler B, Michalski N, Nowossadeck E, Diercke M, Wahrendorf M, Santos-Hövener C, et al. Sozioökonomische Ungleichheit und COVID-19 Eine Übersicht über den internationalen Forschungsstand. Journal of Health Monitoring. 2020;5(S7)(2):1–120. Available from: https://www.rki.de/DE/Content/Gesundheitsmonitoring/Gesundheitsberichterstattung/GBEDownloadsJ/JoHM_2017_02_Gesundheitsverhalten.pdf?__blob=publicationFile.

[26] Yan P, Chowell G. Quantitative Methods for Investigating Infectious Disease Outbreaks. vol. 70 of Texts in Applied Mathematics. Cham: Springer International Publishing; 2019. Available from: http://link.springer.com/10.1007/978-3-030-21923-9.

[27] Bell A, Fairbrother M, Jones K. Fixed and random effects models: making an informed choice. Quality and Quantity. 2019;53(2):1051–1074. Available from: https://doi.org/10.1007/s11135-018-0802-x.

[28] Bell A, Jones K. Explaining Fixed Effects: Random Effects Modeling of Time-Series Cross-Sectional and Panel Data. Political Science Research and Methods. 2015;3(01):133–153.

[29] McCullagh P, Nelder JA. Generalized Linear Models. Second edi ed. Springer Science Business Media, B.V.; 1989.

[30] Harris T, Yang Z, Hardin JW. Modeling underdispersed count data with generalized Poisson regression. Stata Journal. 2012;12(4):736–747.

[31] Public Health England. Investigation of novel SARS-COV-2 variant: Variant of Concern 202012/01 Technical Briefing 3. GovUk. 2020;(December):1–11. Available from: https://www.gov.uk/government/publications/investigation-of-novel-sars-cov-2-variant-variant-of-concern-20201201.

[32] Autorengruppe Bildungsberichterstattung. Bildung in Deutschland 2020. 2020;p. 1–361. Available from: https://www.bmbf.de/de/bildung-in-deutschland-2020-11888.html.

